# Toxic heavy metals in the breast milk of diabetic and non-diabetic postpartum mothers in Yenagoa, Nigeria

**DOI:** 10.1101/2022.02.16.22271066

**Authors:** Tuboseiyefah Perekebi Philip-Slaboh, Chinemerem Eleke, Anthonet Ndidiamaka Ezejiofor

## Abstract

**Background:** Breast milk is considered to be the best substance for neonatal nutrition. It is not well known whether diabetes increases the expression of toxic heavy metals in the breast milk of postpartum mothers. This study compared the concentration of toxic heavy metals in breast milk between diabetic and non-diabetic postpartum mothers in Yenagoa.

**Material and methods:** A cross-sectional design was utilized on a purposive sample of 144 consenting postpartum mothers (72 diabetic and 72 non-diabetic mothers) from one tertiary, one secondary, and one primary medical centre. Breast milk samples were collected at 5-6 weeks postpartum between 1st November 2020 and 30th April 2021. A spectrophotometer was used to analyze the breast milk samples. A pro forma was used for data extraction and data were analyzed at a 5% significance level.

**Result:** The Diabetes group had levels of Arsenic (63.9%), Lead (95.8%), Mercury (68.1%), and Cadmium (84.7%) above the WHO permissible limits. The mean concentrations were 0.6 ng/ml (Arsenic), 13.2ng/ml (Lead), 2.9ng/ml (Mercury), and 3.3ng/ml (Cadmium). The non-diabetic mothers also had high levels of Arsenic (62.5%), Lead (95.8%), Mercury (72.2%), and Cadmium (86.1%); and the mean concentrations were 0.6ng/ml (Arsenic), 12.2 ng/ml (Lead), 3.0ng/ml (Mercury), and 3.2ng/ml (Cadmium). There was no significant difference in the concentration of toxic heavy metals in breast milk between the diabetic and non-diabetic postpartum mothers (p = > 0.585).

**Conclusions:** Diabetes did not seem to increase the concentration of toxic heavy metals expressed in breast milk. More rigorous studies are needed to confirm these findings.

## Introduction

Typically, the postpartum period begins soon after birth and lasts approximately six weeks. Within this period, maternal breast milk serves as the newborn’s only source of nutrition. In its uncontaminated natural state, it contains about 87% of water, 7% of carbohydrates, 4% of lipids, and 1% of proteins, vitamins, and minerals [1]. A disease such as diabetes could alter the electrolyte composition of the human body, and electrolyte deficiency states could mobilize ingested heavy metals into breast milk [2]. The presence of heavy metals in breast milk does present toxicological fears concerning neonatal health [3]. In line with this, the concentration of toxic heavy metals in breast milk requires monitoring especially in regions where environmental pollution is common.

Environmental pollution is linked with one in every six global deaths, and this amounts to about nine million deaths annually [4]. In 2015, polluted air accounted for roughly six million deaths globally [5]. In the same year, water pollution was blamed for approximately two million deaths [6]. Unfortunately, 92% percent of pollution-related deaths occur in low and middle-income countries of the world [4]. Since the growth of global industrialization and fossil fuel exploration, toxic heavy metals such as Arsenic, Lead, Mercury, and Cadmium have progressively been detected in the environment. They make their way into the air from factory and crude-oil refinery emissions and into the water from agriculture and mining industry effluents [7]. Some researchers have implied that when humans are exposed to environments contaminated with high concentrations of heavy metals, these metals can accumulate in human tissue over time. Infants who depend largely on the postpartum mothers’ breast milk may also be at risk of ingesting the named toxic heavy metals from their mothers.

Toxicity from arsenic is currently a public health concern [8]. When arsenic is ingested, it causes skin hypo-pigmentation, peripheral vascular diseases such as black foot disease, gangrene, and high blood pressure [9]. In pregnant women, the chance of cancer is increased and it stimulates growth delay in foetuses. Its presence as a contaminant in breast milk has also been reported [10]. Lead is a heavy metal that naturally exists in rocks. It has no known nutritional value. Contaminated food, water, and air are all sources of lead exposure in humans [11]. In this regard, residential locations near waste yards may have contaminated air, soil, and water. Previous data indicate that when calcium levels in breast milk are low, lead stored in a woman’s bone marrow is physiologically mobilized to match the need [2]. Furthermore, the half-life of lead in human blood is 30 days, whereas the half-life of lead in bone is 27 years. Based on the data, it appears that a breastfeeding mother will continue to shed lead for several days after the initial exposure. Mercury is a heavy element that occurs naturally in the earth’s crust. It is frequently emitted into the environment as a fume by insecticides, herbicides, and battery leaks [12]. Evidence has shown that fish accumulates up to 15 folds more mercury than lead [2]. In line with the mentioned, mercury is ultimately ingested by fish eaters. Cadmium enters the soil and running water through the improper disposal of sewage and computer waste [13]. Literature shows that high quantities of cadmium have been found in oysters, crabs, and the liver and kidneys of some farm animals [11]. Cadmium accumulates in the muscles over a lifetime in humans due to its delayed renal clearance and a half-life of up to 38 years. Evidence suggests that iron deficiency is linked with higher intestinal absorption of cadmium [2]. The postpartum period as a period of low iron stores puts the breastfeeding mother at even greater risk.

Yenagoa is the seat of crude oil exploration in the Niger Delta region. It has experienced several crude oil spills from 1976 till recent times resulting in environmental degradation and the possible release of heavy metals into the environment [14]. The research team wonders if maternal diabetes would worsen the expression of toxic heavy metals in breast milk. A literature search in the major research databases such as MEDLINE, PubMed, and ProQuest yielded few peer-reviewed publications on the subject matter. The knowledge gap motivated the research team to embark on a study to compare the concentration of heavy metals in the breast milk of diabetic and non-diabetic postpartum mothers in Yenagoa, Bayelsa State, Nigeria.

## Material and methods

### Ethics statement

This study was approved by the University of Port Harcourt Institutional Review Board (Approval ID: UPH/CEREMAD/REC/MM75/050) and adhered to the Helsinki Declaration of 1975, as revised in 1983. All participants in this study gave their written consent.

### Study area and period

This study was conducted in three randomly selected postnatal clinics of one tertiary medical centre, one secondary hospital, and one primary medical clinic from 1st November 2020 to 30th April 2021. The facilities were Federal Medical Centre, Diete-Koki Memorial Hospital, and Women Affairs Clinic, all in Yenagoa. Yenagoa is the capital of Bayelsa State and is located deep in the Niger Delta region, in the southern part of Nigeria. It lies between Latitude 4°15^’^ North and Latitude 5°23’ South, and between Longitude 5°22’ West and 6°45’ East. It possesses commercial quantities of crude oil deposits and has about 700,000 residents whose major occupation is fishing and farming. The humongous crude oil exploration activities by multinational companies alongside some clandestine crude oil refining by unlicensed persons make Yenagoa prone to environmental pollution from crude oil spills [15]. The consequent pollution of air, soil, and water raises the potential for human ingestion of toxic heavy metals.

### Study design and population

A cross-sectional design was employed for this study. The study population was an estimated 1,843 postpartum mothers aged between 15 and 49 years. This estimated figure was arrived at by summing up all registered postpartum mothers who obtained postnatal services from the facilities in the year 2019.

### Sample size determination and sampling procedure

A sample size of 144 postpartum mothers (72 diabetic and 72 non-diabetic mothers) was determined for this study using the Cochran’s formula mathematically stated as n = Z^2^ P(1-P)÷d^2^ [16]; where n = minimum sample size; Z = critical value constant 1.96; P = fertility rate for Bayelsa State 4.4% [17]; d = error level 5%. A minimum sample size of 65 was computed. To guard against the threat of fallout, the minimum sample size was increased by 10% using the non-response formula mathematically stated as n* = n ÷ (1-0.1) [18]; where n* = final sample size and n = minimum sample size of 65. A final sample size of 72 was computed for each of the two arms of the study. The purposive sampling technique was applied in the selection and enrolment of consenting participants based on health status and being at 5-6 weeks into the postpartum period.

### Instrument and data collection

A novel nine-item semi-structured data extraction sheet (pro forma) with an inter-rater reliability of 0.938 was used for this study. It had two sections (A and B). Section A was designed to extract the background demographic characteristics of the participants such as age, marital, parity, employment, and disease status. Section B was designed to extract the result of breast milk laboratory analysis for toxic heavy metal composition and concentration. For the collection of breast milk samples, 144 acid-washed 10ml specimen collection plastic bottles were used. A small size cooler box with Ice packs was used for transporting the collected breast milk samples to the chemistry laboratory for analysis. Data and breast milk sample collection took place between November 2020 and April 2021. The purpose of this study was explained to the selected participants. The research team encouraged the consenting participants to give data on their background characteristics using the pro forma (section A). A unique number was coded onto each pro forma and matched with the labels on the acid-washed specimen collection bottles. The participant was encouraged to manually squeeze 10ml of breast milk into the specimen collection bottles and the bottles were put into a small size plastic cooler, packed with ice, and transported for further laboratory analysis in the chemistry laboratory of Ohemerge Company Limited in Port Harcourt, Rivers State. The breast milk samples were tested for heavy metals by the Laboratory Scientist using Atomic Absorption Spectrophotometer (AAS, the AA500 PG model).

### Data analysis

Collected categorical data from participants’ demographic characteristics were summarized using descriptive statistical methods such as frequency and percentage. Interval data from age were summarized using mean, standard deviation, frequency, and percentage. Test of statistical difference between groups was tested using the t-test, Chi-square, and Fisher exact test inferential statistics at a 5% level of significance. All statistical analysis was done with the aid of Statistical Products and Service Solutions software version 25 (IBM Chicago IL, USA).

## Result

Table 1 summarizes the background characteristics of the participants and it revealed no significant difference between the diabetes and non diabetes groups (p= > 0.05). Table 2 summarizes the concentrations of toxic heavy metals in the breast milk of diabetic postpartum mothers and revealed that the majority of the participants in the Diabetic group had high levels of Arsenic (63.9%), Lead (95.8%), Mercury (68.1%), and Cadmium (84.7), above the WHO permissible limits. The mean concentrations compared with the WHO permissible limits (Reference) of the toxic heavy metals in the sample were Arsenic (0.6 vs. 0.0 ng/ml), Lead (13.2 vs. 5.0 ng/ml), Mercury (2.9 vs. 1.7 ng/ml) and Cadmium (3.3 vs. 1.0 ng/ml). Table 3 summarizes the concentration of toxic heavy metals in the breast milk of the Non-diabetic group and revealed that the majority of the participants had high levels of Arsenic (62.5%), Lead (95.8%), Mercury (72.2%), and Cadmium (86.1), above the WHO permissible limits. The mean concentrations compared with the WHO permissible limits of the toxic heavy metals in the sample were Arsenic (0.6 vs. 0.0 ng/ml), Lead (12.2 vs. 5.0 ng/ml), Mercury (3.0 vs. 1.7 ng/ml), and Cadmium (3.2 vs. 1.0 ng/ml). Table 4 compares toxic heavy metal concentrations in the breast milk of Diabetic and Non-diabetic groups and revealed no significant difference between the groups (p > 0.05).

**Table 1.**
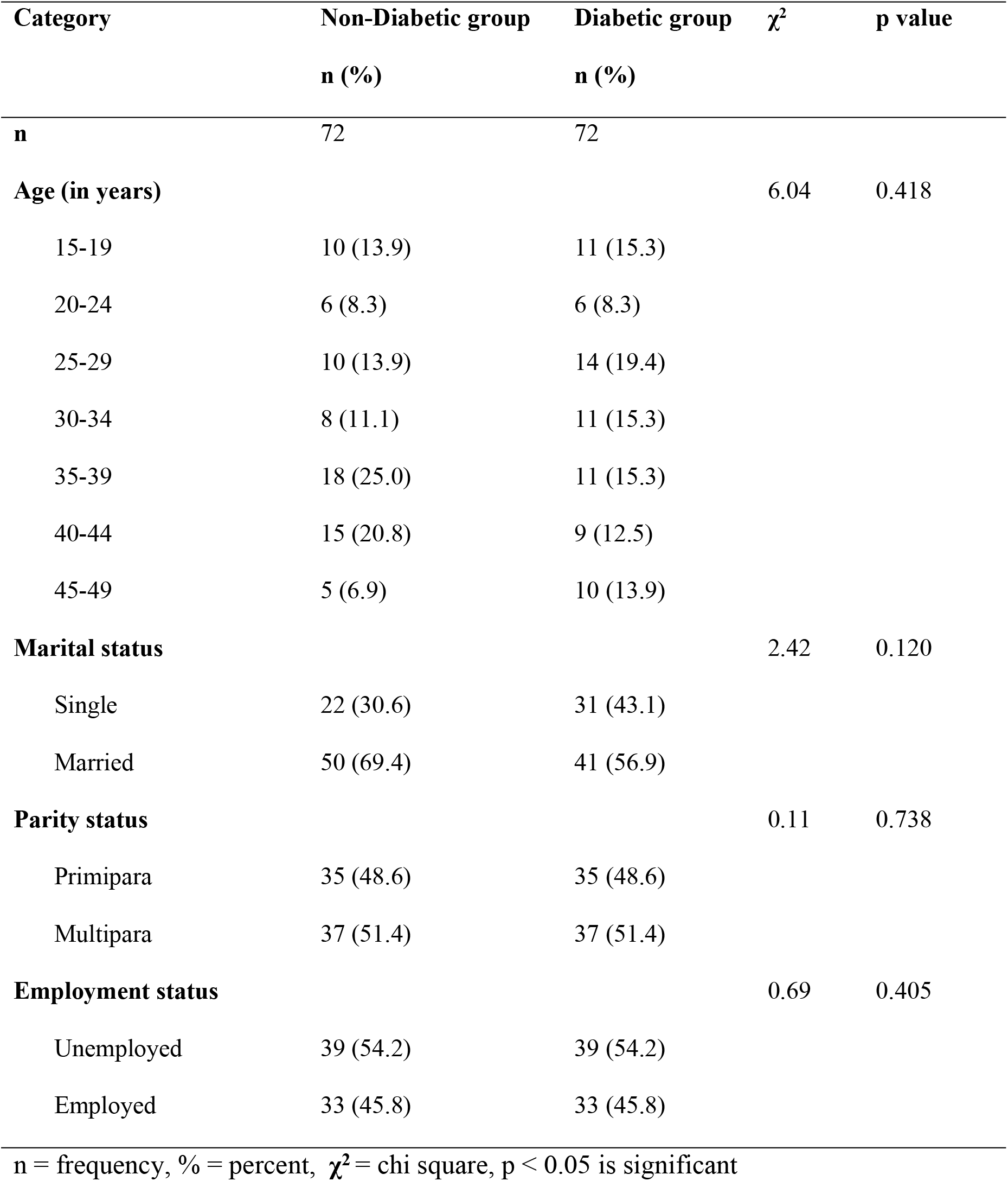
Background characteristics of the participants **N = 144**

**N = 144**
| Category | Non-Diabetic group | Diabetic group | $\chi^2$ | p value |
| --- | --- | --- | --- | --- |
|  | n (%) | n (%) |  |  |
| **n** | 72 | 72 |  |  |
| **Age (in years)** |  |  | 6.04 | 0.418 |
| 15-19 | 10 (13.9) | 11 (15.3) |  |  |
| 20-24 | 6 (8.3) | 6 (8.3) |  |  |
| 25-29 | 10 (13.9) | 14 (19.4) |  |  |
| 30-34 | 8 (11.1) | 11 (15.3) |  |  |
| 35-39 | 18 (25.0) | 11 (15.3) |  |  |
| 40-44 | 15 (20.8) | 9 (12.5) |  |  |
| 45-49 | 5 (6.9) | 10 (13.9) |  |  |
| **Marital status** |  |  | 2.42 | 0.120 |
| Single | 22 (30.6) | 31 (43.1) |  |  |
| Married | 50 (69.4) | 41 (56.9) |  |  |
| **Parity status** |  |  | 0.11 | 0.738 |
| Primipara | 35 (48.6) | 35 (48.6) |  |  |
| Multipara | 37 (51.4) | 37 (51.4) |  |  |
| **Employment status** |  |  | 0.69 | 0.405 |
| Unemployed | 39 (54.2) | 39 (54.2) |  |  |
| Employed | 33 (45.8) | 33 (45.8) |  |  |

n = frequency, % = percent, $\chi^2$ = chi square, p < 0.05 is significant

**Table 2.** Concentration of toxic heavy metals in the breast milk of the Diabetic group **N = 72**

| Parameters | Arsenic | Lead | Mercury | Cadmium |
| --- | --- | --- | --- | --- |
| Normal level n (%) | 26 (36.1) | 3 (4.2) | 23 (31.9) | 11 (15.3) |
| High level n (%) | 46 (63.9) | 69 (95.8) | 49 (68.1) | 61 (84.7) |
| Mean (SD) ng/ml | 0.63 (0.36) | 13.20 (5.13) | 2.98 (1.91) | 3.32 (1.86) |
| Minimum ng/ml | 0.0 | 4.3 | 0.1 | 0.1 |
| Maximum ng/ml | 1.2 | 22.4 | 6.3 | 6.3 |
| WHO limits ng/ml (Reference) | 0.0 | 5.0 | 1.7 | 1.0 |

n = frequency, % = percent, SD = standard deviation, ng/ml = nanogram per mililitre

**Table 3.**
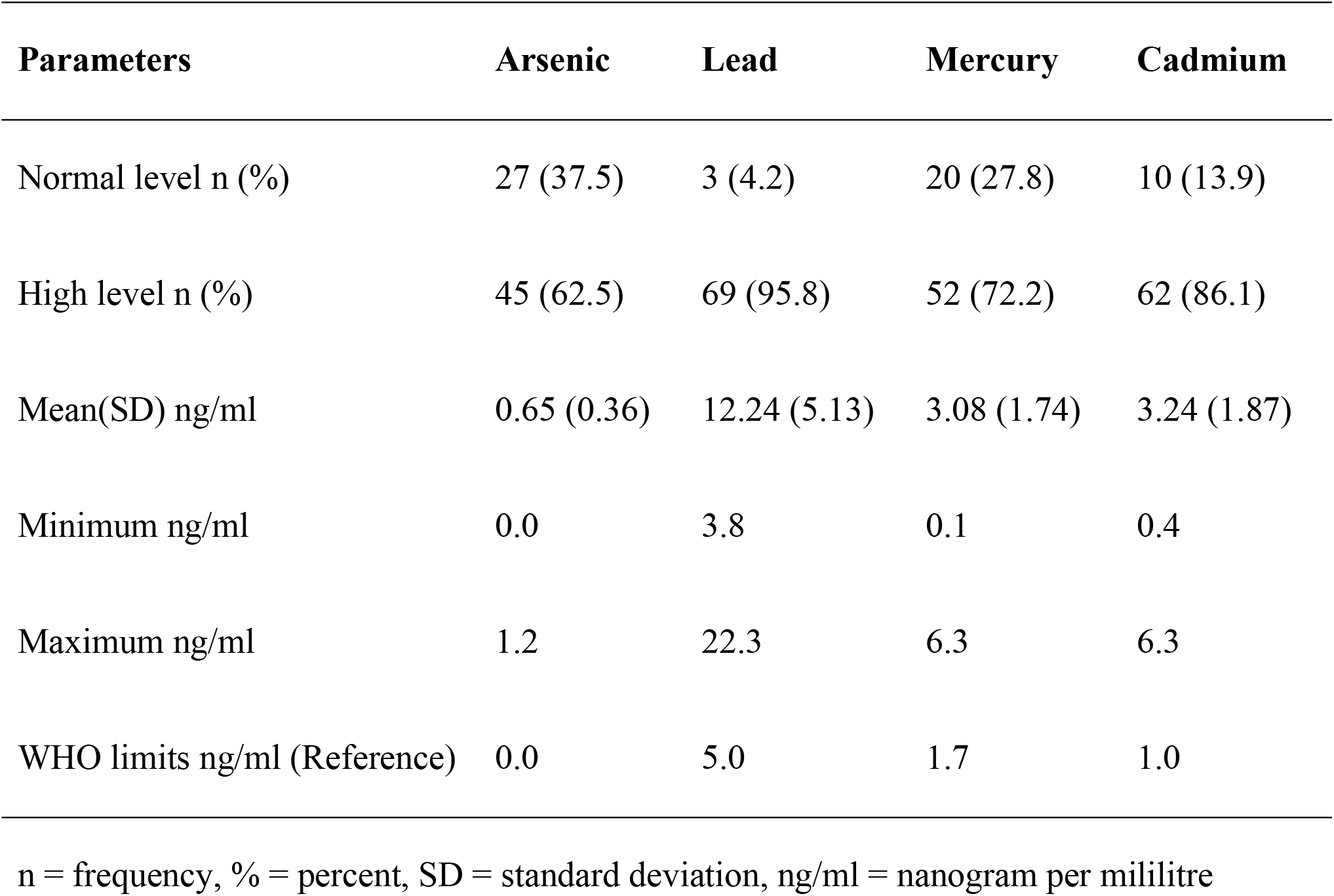
Concentration of toxic heavy metals in the breast milk of the Non-diabetic group **N = 72**

| **Parameters** | **Arsenic** | **Lead** | **Mercury** | **Cadmium** |
| --- | --- | --- | --- | --- |
| Normal level n (%) | 27 (37.5) | 3 (4.2) | 20 (27.8) | 10 (13.9) |
| High level n (%) | 45 (62.5) | 69 (95.8) | 52 (72.2) | 62 (86.1) |
| Mean(SD) ng/ml | 0.65 (0.36) | 12.24 (5.13) | 3.08 (1.74) | 3.24 (1.87) |
| Minimum ng/ml | 0.0 | 3.8 | 0.1 | 0.4 |
| Maximum ng/ml | 1.2 | 22.3 | 6.3 | 6.3 |
| WHO limits ng/ml (Reference) | 0.0 | 5.0 | 1.7 | 1.0 |

n = frequency, % = percent, SD = standard deviation, ng/ml = nanogram per mililitre

**Table 4.** Comparison of toxic heavy metal concentrations in the breast milk of Diabetic and Non-diabetic groups **N = 144**

| **Variable** | **Diabetes group**<br>**n** | **Non-diabetic group**<br>**n** | **t-test** | **$\chi^2$** | **P value** |
| --- | --- | --- | --- | --- | --- |
| **Arsenic** Mean(SD) | 0.63 (0.36) | 0.65 (0.36) | 0.39 |  | 0.696 |
| Normal level | 26 | 27 |  | 0.03 | 0.863 |
| High level | 46 | 45 |  |  |  |
| **Lead** Mean(SD) | 13.20 (5.14) | 12.24 (5.13) | -1.12 |  | 0.263 |
| Normal level | 3 | 3 |  | 0.00 | 1.000 |
| High level | 69 | 69 |  |  |  |
| **Mercury** Mean(SD) | 2.90 (1.91) | 3.08 (1.74) | 0.31 |  | 0.757 |
| Normal level | 23 | 20 |  | 0.30 | 0.585 |
| High level | 49 | 52 |  |  |  |
| **Cadmium** Mean(SD) | 3.32 (1.86) | 3.24 (1.87) | -0.25 |  | 0.803 |
| Normal level | 11 | 10 |  | 0.06 | 0.813 |
| High level | 61 | 62 |  |  |  |

n = frequency, SD = standard deviation, Mean(SD) was meaasured in ng/ml, p < 0.05 is significant

## Discussion

This study found that majority of the participants in the Diabetes group had high concentrations of Arsenic, Lead, Mercury, and Cadmium in their breast milk (higher than the WHO permissible limits). This finding partially corroborates an Egyptian study that found that Lead (2,920 vs.5.0 ng/ml) and Cadmium (16800 vs. 1.0ng/ml) at toxic concentrations higher than the WHO permissible limits, but did not detect Arsenic and Mercury in the breast milk samples in Damietta [19]. Moreover, the concentrations of Lead and Cadmium that were found in the Egyptian study were about 200 folds more for Lead and 500 folds more for Cadmium. The dissimilarity in findings is connected to the activities predominant in the area of study. Diametta was reported to be an agrarian society that uses compost as fertilizers and has a high factory and vehicular traffic emissions. Compost is known to increase the soil composition of Copper, Zinc, and Lead. Additionally, petrol and gas cars are likely to librate even more Lead while factories will emit cadmium in exhaust fumes. In contrast, the area of this present study is not known for its use of compost as fertilizer but crops have been reportedly destroyed over the years due to environmental crude oil spills [14]. Due to the disparity in the qualities of the studied areas, the dissimilarity in findings between the studies was expected. Also, this finding was lower than that reported in a study conducted in Cyprus that found Arsenic (730 ng/ml vs. 0.0 ng/ml), Lead (1,190 vs. 5.0ng/ml), and Cadmium (450 vs. 1.0 ng/ml) at very high amounts in breast milk samples. The discrepancy in findings could be linked to the inequalities in sample attributes utilized in the study [20]. The Cyprian study examined breast milk samples that were drawn from a mix of urban and rural postpartum women hence a heterogeneous sample. A heterogeneous sample limits the internal validity of the study. In contrast, this present study examined breast milk samples drawn from a homogenous urban population. In the light of the fore mentioned, the findings between this study and the Cyprian study were expected to differ.

This study found that the majority of the participants in the Non-diabetic group had high levels of Arsenic, Lead, Mercury, and Cadmium in their breast milk samples (above the WHO permissible limits). This finding corroborates a Zanzibarian study which reported that Lead (707 vs.5.0 ng/ml) and Cadmium (311 vs. 1ng/ml) were found in breast milk samples at levels that were greater than the WHO set limits [21]. Likewise, the Lead and Cadmium levels reported in the Zanzibarian study were greater than what was found in this study by 25 and 100 folds respectively. The dissimilarity in findings is connected to the points in the postpartum period when the breast milk samples were collected for examination. The Zanzibarian study collected breast milk samples from participants irrespective of the number of weeks into the postpartum period that the participants were. Based on the context that the expression of toxic heavy metals is more expressed in the colostrum than in mature milk, a mix of breast milk samples at different stages of lactation limits the internal validity of the study [3]. Breast milk samples obtained from women at more than 6 weeks postpartum were likely to provide findings that were not a fair representation of the snapshot of the phenomena of interest due to data set contamination. In this present study, the researcher specifically collected breast-milk samples at 5-6 weeks into the postpartum period. It was done to provide a more valid snapshot of the concentrations of the toxic heavy metals at one specific point in time across the sample examined. The dissimilarity in findings between this study and the Zanzibarian study was hence expected. Likewise, this finding did not completely support another Iranian study that reported that Lead (7.15 vs. 5.0 ng/ml) and Cadmium (1.07 vs. 1ng/ml) were detected in breast milk and Lead was notably above the WHO preset limit [22]. The similarity in findings could be connected to the design utilized in the study. Both the Iranian study and the present study utilized the cross-sectional design and without manipulation of variables. However, the levels reported in the Iranian study were lower than the values noted in this present study. The discrepancy in value could be linked to the method of laboratory examination of the breast milk samples utilized. The Iranian study utilized the anodic stripping voltammetry method for the laboratory testing of the breast milk samples, whereas this present study utilized the Atomic Absorption Spectrophotometry method. Some probably inherent disparity in processing and analysis between the two methods may have resulted in the discrepancy in concentrations.

This study found no statistically significant difference in the level of toxic heavy metals in breast milk between the Diabetes and Non diabetes groups. This finding may imply that diabetes has no substantial impact on the expression of toxic heavy metals in their breast milk. The protein in breast milk is known to have a high potential for heavy metal binding [23]. Mature milk has less protein than Colostrum but more fat and sugar, and the high sugar levels start to decrease at approximately 18 months after birth [24]. It thus implies that as human breast milk matures into the 5-6 weeks after childbirth when fewer proteins and immunoglobulins are expressed in breast milk, the amount of heavy metals bound to the protein expressed in breast milk thus reduces back to near-normal levels [3]. In line with this, similarities in the level of toxic heavy metals in the breast milk of diabetic postpartum mothers should be similar to those of non-diabetic postpartum mothers.

A notable limitation to this study is that it did not assess maternal involvement in clandestine refining of crude oil which involves direct contact with possible toxicants. It was omitted from the proforma for data gathering because of ethical issues, as clandestine crude oil refining is considered a social crime as artisanal refineries are neither registered nor licensed by the government. The participants were foreseen as not willing to provide such information for fear of arrest and litigation. Additionally, this study utilized the non-probability-based purposive sampling method. Non-probability-based sampling techniques do not offer an equal chance of selection and enrollment to all members of a target population. This, therefore, limited the data set’s capacity to meet the statistical criteria for more robust parametric statistical tests. These possible uncertainties perhaps impose a fair chance of committing type II error. Until further risk assessment studies with bigger samples become available, the generalization of these findings outside of this study group should be done with caution.

## Conclusion

The study participants had an undesirably high concentration of toxic heavy metals in their breast milk but disease status did not result in any significant differences between the diabetic and Non-diabetic mothers at five to six weeks following childbirth. More toxicological research on mother and child health may be necessary to validate the conclusions of this study.

## Data Availability

All relevant data are within the manuscript and its Supporting Information files.

## Acknowledgements

We thank the staff of Ohemerge Company Limited Port Harcourt, Rivers State for allowing us use their laboratory facility for the chemical analysis of breast milk samples.

## References

1. Pajewska-Szmyt M, Sinkiewicz-Darol E, & Gadzała-Kopciuch R. The impact of environmental pollution on the quality of mother’s milk. Environ Sci Pollut Res Int. 2019; 26(1): 7405–27. https://doi.org/10.1007/s11356-019-04141-1.

2. Yurdakok K. Lead, mecury, and cadmium in breast milk. J Pediatr Neonat Individual Med. 2015; 4(2): Article ID e040223. https://doi.org/10.7363/040223.

3. Chao H-H, Guo C-H, Huang C-B, Chen P-C, Li H-C, Hsiung D-Y, et al. Arsenic, cadmium, lead, and aluminium concentrations in human milk at early stages of lactation. Pediatr Neonatol. 2014; 55(2): 127–34. https://doi.org/10.1016/j.pedneo.2013.08.005.

4. Mayor S. Pollution is linked to one in six deaths worldwide, study estimates. BMJ. 2017; 359(1): Article ID j4844. https://doi.org/10.1136/bmj.j4844.

5. Landrigan P. Pollution and health. Lancet Public Health. 2017; 2(1): Article ID e23. https://doi.org/10.1016/S2468-2667(16)30023-8.

6. Prüss-Üstun A, Wolf J, Corvalán C, Bos R, Neira, M. Preventing disease through healthy environments: a global assessment of the burden of disease from environmental risks. Geneva: World Health Organization; 2016.

7. Obasi P, Akudinobi, B. Potential health risk and levels of heavy metals in water resources of lead–zinc mining communities of Abakaliki, southeast Nigeria. Appl Water Sci. 2020; 10(1): Article ID184. https://doi.org/10.1007/s13201-020-01233-z.

8. Wu H, Wang M, Raman J, McDonald A. Association between urinary arsenic, blood cadmium, blood lead, and blood mercury levels and serum prostate-specific antigen in a population-based cohort of men in the United States. PLoS One. 2021; 16(4): Article ID e0250744. https://doi.org/10.1371/journal.pone.0250744.

9. Briffa J, Sinagra E, Biundell R. Heavy metal pollution in the environment and their toxicological effects on humans. Heliyon. 2020; 6(9): Article ID e0491. https://doi.org/10.1016/j.heliyon.2020.e04691.

10. Hameed A, Akhtar S, Amjad A, Naeem I, Tariq M. Comparative assessment of arsenic contamination in raw milk, infant formulas and breast milk. J Dairy Vet Sci. 2019; 13(1): Article ID 555851. https://doi.org/10.19080/JDVS.2019.13.555851.

11. Nkwunonwo U, Odika P, Onyia NI. (2020). A review of the health implications of heavy metals in food chain in Nigeria. ScientificWorldJournal. Article ID 6594109. https://doi.org/10.1155/2020/6594109.

12. Rehman K, Fatima F, Waheed I, Akash M. Prevalence of exposure of heavy metals and their impact on health consequences. J Cell Biochem. 2017; 119(1): 157–84. https://doi.org/10.1002/jcb.26234.

13. Winiarska-Mieczan A. Cadmium, lead, copper and Zinc in breast milk in Poland. Biol Trace Elem Res. 2014; 157(1): 36–44. https://doi.org/10.1007/s12011-013-9870-x.

14. Chinedu E, Chukwuemeka C. Oil spillage and heavy metals toxicity risk in the Niger Delta, Nigeria. J Health Pollut. 2018; 8(19): Article ID 180905. https://doi.org/10.5696/2156-9614-8.19.180905.

15. Ibegu K, Olusola A. Effects of oil operations on Epebu community in Bayelsa State. Convent J Res Built Environ. 2017; 5(2): 58–74.

16. Charan J, Biswas, T. How to calculate sample size for different study designs in medical research. Indian J Psychol Med. 2013; 35(2): 121–6. https://doi.org/10.4103/0253-7176.116232.

17. National Population Commission & ICF-World Health Organization. Demographic and health survey 2018. Abuja: NPC and ICF; 2019.

18. Bolarinwa O. Sample size estimation for health and social science researchers: The principles and considerations for different study designs. Niger Postgrad Med J. 2020; 27(1): 67–75.

19. Hasballah A, Beheary M. Detection of heavy metals in breast milk and drinking water in Damietta Governorate, Egypt. Asian J Biol. 2016; 1(2): 1–7. https://doi.org/10.9734/AJOB/2016/30517.

20. Kunter I, Hurer N, Gulcan H, Ozturk B, Dogan I, Sahin G. Assessment of aflatoxin M1 and heavy metal Levels in mothers breast milk in Famagusta, Cyprus. Biol Trace Elem Res. 2017; 175(1): 42–9. https://doi.org/10.1007/s12011-016-0750-z.

21. Khamis H, Lusweti K, Mwevura H. Quantification of heavy metals in breast milk samples sampled from Kilimani/Kidoti in Zanzibar. American Scientific Res J Engineering Technology Sci. 2017; 35(1): 295–308.

22. Sadeghi N, Jannat B, Behzad M, Oveisi M, Hajimahmoodi M, Ahmadi F. Determination of zinc, copper, lead and cadmium concentration in breastmilk by anodic stripping voltammetry method and investigating their impact on infants’ growth indicators. J Human Health Halal Metrics. 2020; 1(2): 24–30. https://doi.org/10.30502/JHHHM.2020.226846.1015.

23. Christian P, Smith E, Lee S, Vargas A, Bremer A., Raiten D. The need to study human milk as a biological system. Am J Clin Nutr. 2021; 113(5): 1063–72. https://doi.org/10.1093/ajcn/nqab075.

24. Awua A, Boatin R, Adom T, Brown-Appiah E, Diaba A, Datohe D, et al. Double burden of malnutrition: Toxic metals in breast milk may limit the amounts of micronutrients available to infants through breast milk. Food Nutr Sci. 2019; 10(1); 298–314. https://doi.org/10.4236/fns.2019.103023.

